# ACTIVE SURVEILLANCE OF THE SPUTNIK V VACCINE IN HEALTH WORKERS

**DOI:** 10.1101/2021.02.03.21251071

**Authors:** Vanina Pagotto, Analia Ferloni, María Mercedes Soriano, Morena Díaz, Manuel Braguisnky Golde, María Isabel González, Valeria Asprea, Inés Staneloni, Gustavo Vidal, Martin Silveira, Paula Zingoni, Valeria Aliperti, Hernan Michelangelo, Silvana Figar

## Abstract

**Introduction:** The World Health Organization (WHO) recommends vaccination against Sars Cov-2 coronavirus to mitigate COVID-19 pandemic. On December 29th, the Argentine Ministry of Health started a vaccination plan with the Sputnik V vaccine emphasizing the registration of the Events Supposedly Attributed to Vaccines and Immunizations (ESAVI) in the National Surveillance System. The aim of this study is to determine the safety of this vaccine.

**Methods:** In an ongoing cohort study, health professionals from Hospital Italiano de Buenos Aires vaccinated with the first component of the Sputnik V vaccine (a rAd26 vector-based) were followed up. Safety at 72 hs was analysed from a self-report form. Local and systemic reactions were characterized as mild, moderate and severe. Incident rates were calculated per 1000 person-hours by age groups and gender. Adjusted hazard ratio and 95% Confidence Interval (HR; 95%CI) is obtained by Cox Regression Model.

**Results:** 707 health professionals (mean age 35, 67% female) were vaccinated, response rate was 96,6% and 71,3% reported at least one ESAVI. Rate was 6.3 per 1.000 person-hours. Among local reactions, 54% reported pain at the injection site, 11% redness and swelling. Among systemic reactions 40% reported fever, 5% diarrhea and 68% new or worsened muscle pain. Five percent had serious adverse events that required medical evaluation and one inpatient.

ESAVI rate was higher among females (65.4% vs 50%; HR 1.38, 95%CI 1.13-5.38) and in younger than 55 years-old (72.8% vs 32%; HR 2.66, 95%CI 1.32-1.68).

**Conclusion:** Active surveillance on safety for vaccines with emergency approval is mandatory. This study shows high rates of local and systemic reactions however early serious events were rare. Short term safety is supported by these preliminary findings. Studies on long term safety and efficacy, accoding sex and age, are needed.

## Introduction

The World Health Organization (WHO) recommends vaccination against Sars Cov-2 coronavirus to mitigate COVID-19 pandemic. Effective and safe vaccines in the short term will help to reduce the incidence of illness, hospitalizations and deaths related to COVID-19 and help to gradually restore a new normality in the functioning of our country.^1^

The Sputnik V is a heterologous COVID-19 vaccine consisting of two immunogenic components, a recombinant adenovirus type 26 (rAd26) vector and a recombinant adenovirus type 5 (rAd5) vector, both carrying the gene for severe acute respiratory syndrome coronavirus 2 (SARS-CoV-2) spike glycoprotein (rAd26-S and rAd5-S), it is applied in two doses separated by at least 21 days.^2,9^

On December 23, the National Administration of Medicines, Food and Medical Technology (ANMAT)^3^,10 performed a technical and confidential report on the Sputnik V vaccine; the National Ministry of Health approved it as an “emergency authorization” under the framework of a law 27573, specially approved for this purpose.^3^

The National Ministry of Health^3,4^ developed an Strategic Plan for ensuring the processes to reach the standards of safety and efficacy for the entire Argentine territory, being vaccination in stages, free, voluntary and independent of the history of having suffered the disease.^5^

For the surveillance of vaccine safety, the Strategic Plan proposes health effectors “*To develop a specific plan for intensified passive and active surveillance of vaccine safety, which allows the continuous analysis of the notifications of ESAVI (Events Supposedly Attributed to Vaccines and Immunizations)”*.^5^

Faced with this particular situation of epidemic emergency, which enables the population administration of vaccines without completing phase 3, concurrent safety research with the vaccination campaign is essential. The Ministry of Health of Buenos Aires City (CABA) started, as a public policy, an active surveillance registry of any vaccine applied to health workers.^12 6^ In this uncertain reality, collaborative tools of information and communication technologies (ICTs), like an ESAVI self-report form, are helpful to design collaborative epidemiological studies.

On 12/29/2020 the vaccination campaign started at CABA for those who were health professionals, both from public and private effectors, with an initial endowment of 24,000 Sputnik V vaccine schemes, granted by the National Ministry of Health in 2 batches.

The present study is a preliminary analysis of an ongoing multicenter study of private health institutions from the CABA, to describe the incidence of events supposedly attributed to vaccines in health workers after immunization with the first component of the Sputnik V vaccine.

## Materials and methods

An observational and analytical study of a prospective cohort represented by health workers immunized with the Sputnik V vaccine was carried out at the Hospital Italiano de Buenos Aires. The surveillance period was defined from the date of administration of the first component of the vaccine up to 72 hours after.

A self-report form was sent by email and was forwarded up to three times. Those workers who did not respond to the email were called by a nurse. Those reactions that after medical evaluation were considered ESAVI were reported to the Argentine Integrated Health Information System (SISA). The Ministry of Health of CABA designed an expanded file in the electronic medical record where the clinical monitoring and typification of the ESAVI is recorded, as well as the information prior to vaccination, including the history of COVID-19. Based on this file, in public institutions and some private ones, the Immunization Program of the Ministry of Health of CABA reports centrally to the Argentine Integrated Health Information System.

### Statistical analysis

Quantitative data were expressed as mean and standard deviation or median and interquartile range (IQR) according to distribution. Qualitative data were expressed as absolute and relative frequencies.

The age groups were defined in categories every 10 years (unifying in the same group from 18 to 30 years old) and to allow external comparisons with other publications also in a dichotomous category less than or equal to 55 years old and greater than 55.

The global cumulative incidence of ESAVI was estimated by sex and age groups, considering as denominator the total number of people who responded to the self-report form. The 95% confidence intervals (95% CI) were estimated.

To evaluate the speed of appearance of ESAVI, the incidence was estimated using the Kaplan Meier estimator. The factors associated with the incidence of ESAVI were evaluated using Cox regression. The crude and adjusted Hazard Ratio (HR) were expressed with their 95% CI. A level of statistical significance less than 0.05 was considered. The analysis was carried out with software R version 4.0.3

The protocol was approved by the Research Protocol Ethics Committee of the Hospital Italiano de Buenos Aires under number 3876

## Results

Between January 5^th^ and January 20^th^ 2021, a total of 707 health workers were vaccinated. Six hundred eighty three answered the self-report 683, with a response rate of 96.6%. Of the 683, thirty four workers had had coronavirus detected before the immunitation by real-time polymerase chain reaction. **Table 1** describes the baseline characteristics of the vaccinated health workers.

**Table 1.**
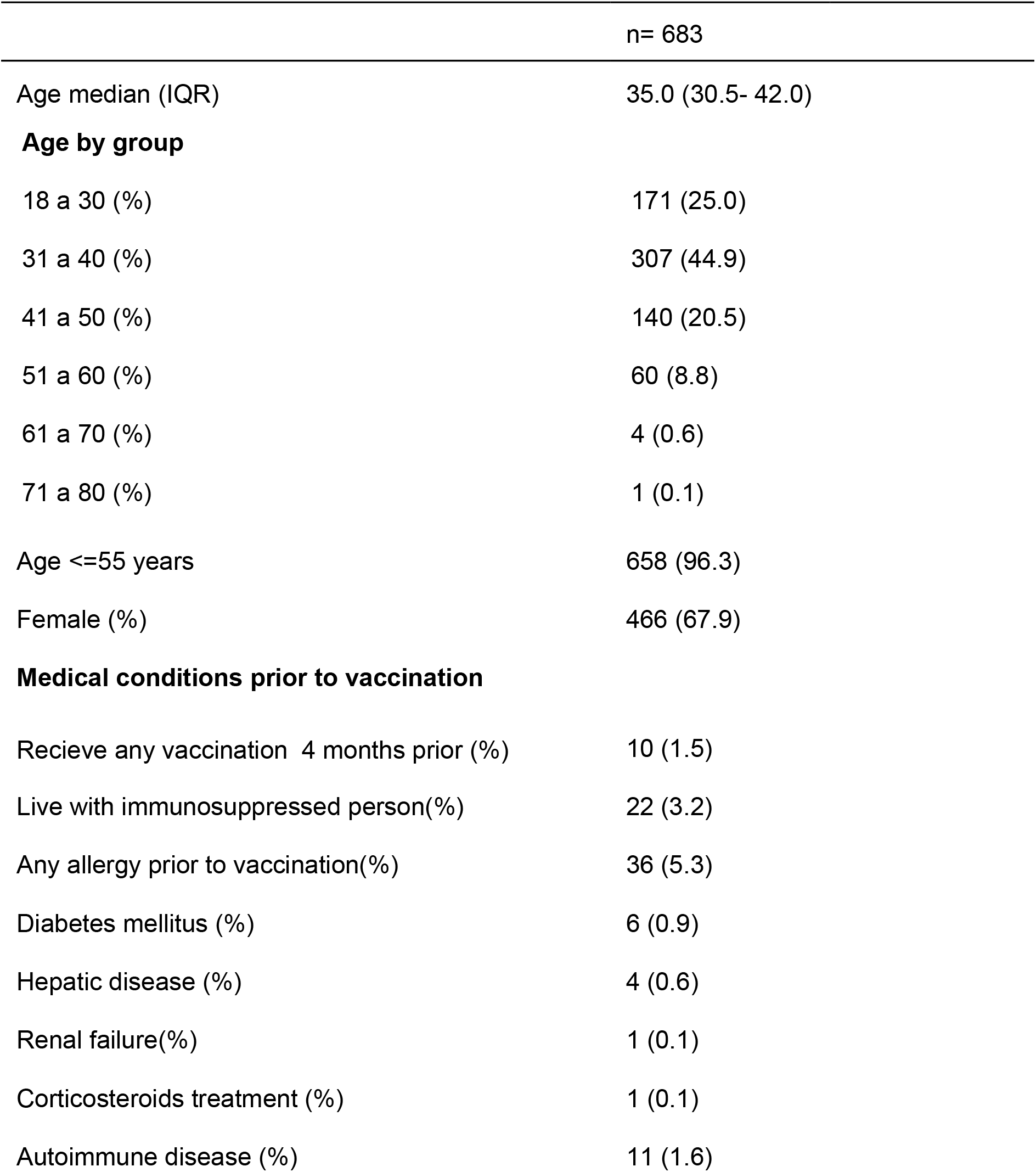
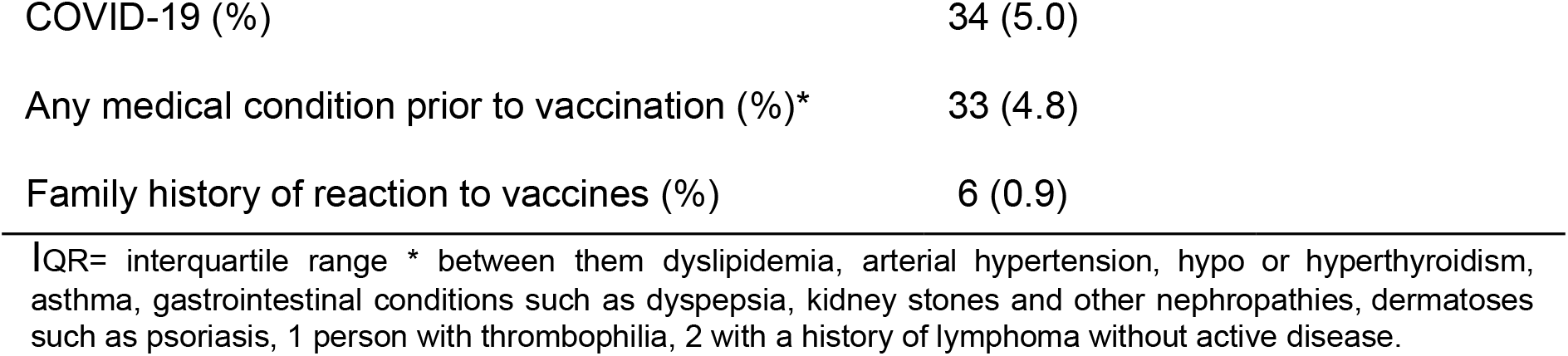
Basal characteristic of vaccinated health workers.

|  | n= 683 |
| --- | --- |
| Age median (IQR) | 35.0 (30.5- 42.0) |
| **Age by group** |  |
| 18 a 30 (%) | 171 (25.0) |
| 31 a 40 (%) | 307 (44.9) |
| 41 a 50 (%) | 140 (20.5) |
| 51 a 60 (%) | 60 (8.8) |
| 61 a 70 (%) | 4 (0.6) |
| 71 a 80 (%) | 1 (0.1) |
| Age <=55 years | 658 (96.3) |
| Female (%) | 466 (67.9) |
| **Medical conditions prior to vaccination** |  |
| Recieve any vaccination 4 months prior (%) | 10 (1.5) |
| Live with immunosuppressed person(%) | 22 (3.2) |
| Any allergy prior to vaccination(%) | 36 (5.3) |
| Diabetes mellitus (%) | 6 (0.9) |
| Hepatic disease (%) | 4 (0.6) |
| Renal failure(%) | 1 (0.1) |
| Corticosteroids treatment (%) | 1 (0.1) |
| Autoimmune disease (%) | 11 (1.6) |
| COVID-19 (%) | 34 (5.0) |
| Any medical condition prior to vaccination (%)* | 33 (4.8) |
| Family history of reaction to vaccines (%) | 6 (0.9) |

IQR= interquartile range \* between them dyslipidemia, arterial hypertension, hypo or hyperthyroidism, asthma, gastrointestinal conditions such as dyspepsia, kidney stones and other nephropathies, dermatoses such as psoriasis, 1 person with thrombophilia, 2 with a history of lymphoma without active disease.

From 683 health workers, 487 (71.3%) had at least one ESAVI. **Figure 1** shows the frequency of ESAVI. No events of special interest were observed (vaccine-augmented disease; multisystemic inflammatory syndrome; respiratory distress; acute heart failure; cardiomyopathy; arrhythmias; coronary artery disease; myocarditis; acute kidney failure; acute liver failure; Guillán Barré; encephalopathy; acute disseminated encephalomyelitis; transverse myelitis; seizures; meningoencephalitis; thromboembolism; thrombocytopenia vasculitis; acute septic arthritis; erythema multiforme; perneum erythema; anaphylaxis)

**Figure 1.**
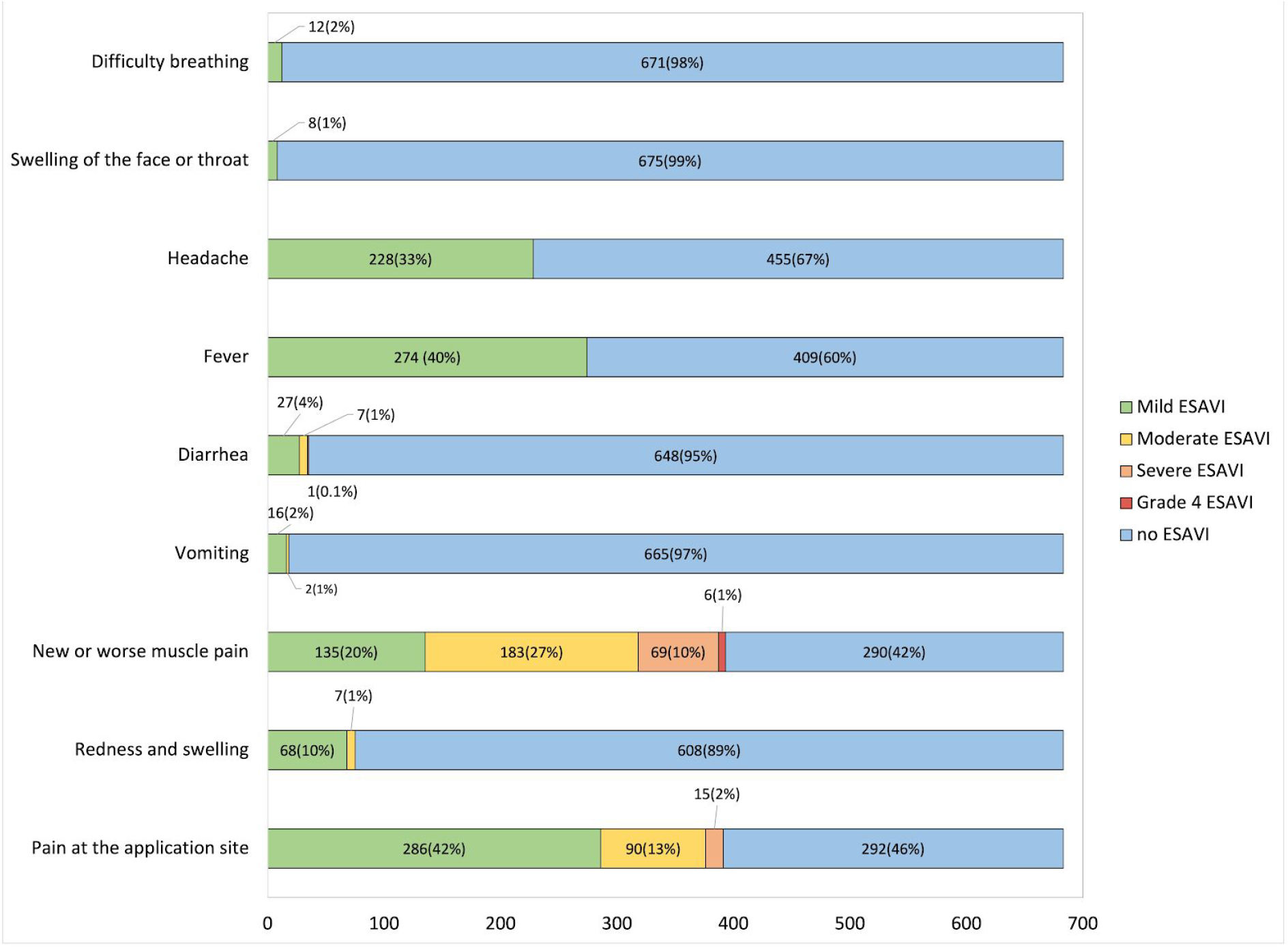
Frequency of Events Supposedly Attributed to Vaccines and Immunizations (ESAVI) The figure shows the axis of the and the type of Events Supposedly Attributed to Vaccines and Immunizations (ESAVI) and within each column the absolute frequency is observed and the relative frequency in percentages in parentheses. The x-axis shows the total number of immunized health workers who responded to the self-report

Regarding the evolution of the ESAVI, 422 (86.6%) had ad-integrum restitution, and 25 (5.1%) needed medical assessment; one person was hospitalized for an acute abdomen that resolved favorably without surgery, and 3 people had COVID-19 diagnosed within 72 hours after vaccination.

When evaluating the incidence of ESAVI by sex and age groups, it was observed that these were more frequent in younger health workers and in women. **Figure 2**

**Figure 2.**
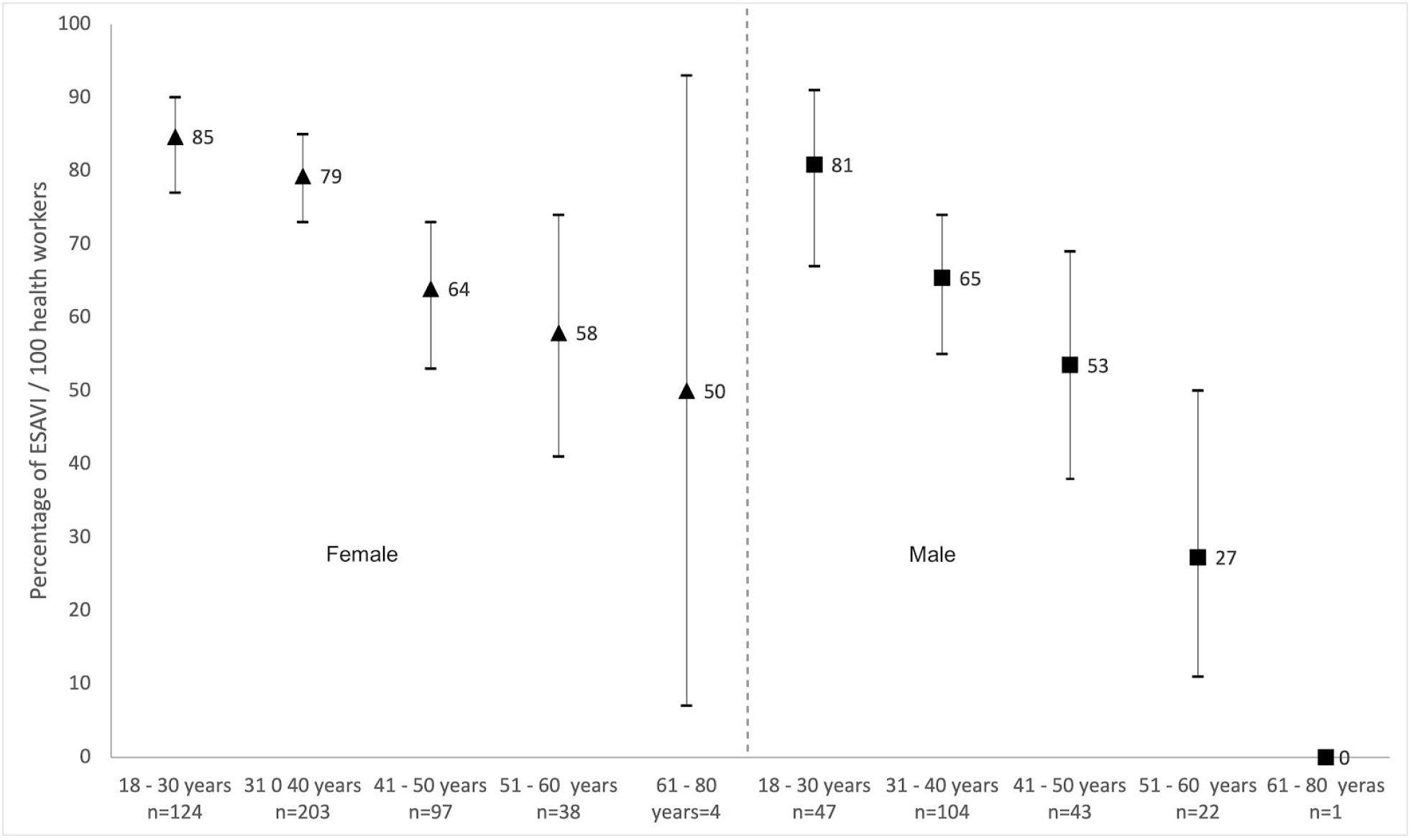
Global incidence of Events Supposedly Attributed to Vaccines and Immunizations (ESAVI) by age group and sex per 100 health workers. The figure shows the incidence of adverse events related to the vaccine. On the x-axis are the age group categories with their absolute frequency (n). On the y axis, the incidence of adverse events per 100 people is observed. Inside the graph, the triangles correspond to the incidence in women and the squares to the incidence in men, the dashes joined by the bars are the upper and lower 95% confidence intervals

Regarding the age groups, less than or equal to 55 years old and greater than 55 years old, the incidence of ESAVI was higher in the first group, being observed in 479 vaccinated (72.8%) and only in 8 (32%) respectively (p <0.001). The speed at which ESAVI appeared is shown by an incidence density of 6.3 per 1,000 patient-hours and the incidence of ESAVI at 48 hours was 60.5% (57.0-64.2).

The cumulative incidence at 48 hours was higher in women (65.4%; 61.1-69.7) than in men (50.0%, 45-58.1%). **Figure 3** shows the incidence of ESAVI in women and men.

**Figure 3.**
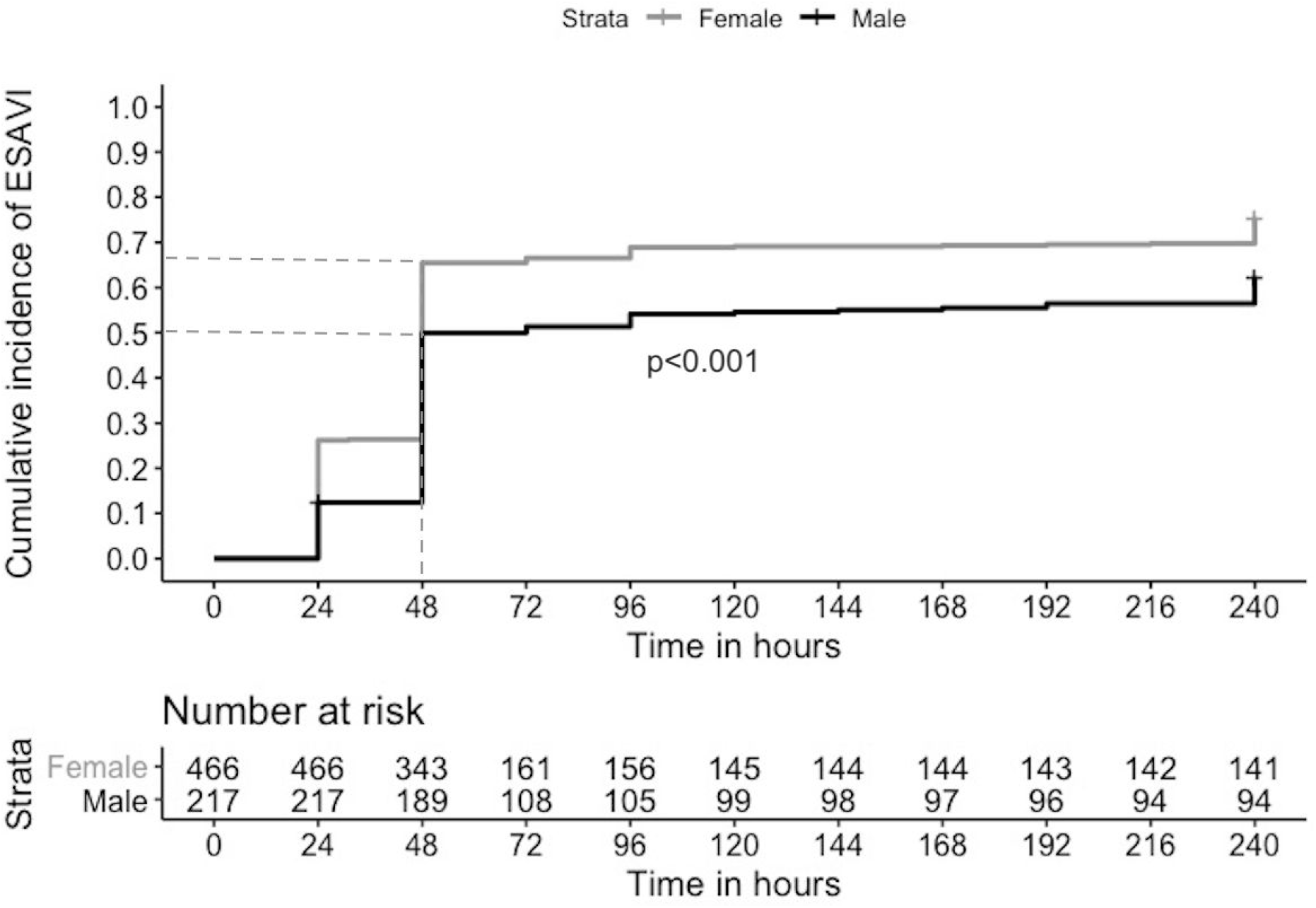
Cumulative incidence of Events Supposedly Attributed to Vaccines and Immunizations (ESAVI) according to sex. Kaplan Meier curve. The dotted line corresponds to the estimated incidence of ESAVI at 48 hours. Number at risk is the number of people at risk every 24 hours from the date of vaccination. The p value compares the curves between men and women.

Likewise, the cumulative incidence of ESAVI at 48 hours was higher in health workers aged up to 55 years (61.8%, 58.1-65.5) than in those over 55 years (28.0%, 14.1-49.9). **Figure 4** shows the incidence of ESAVI in these age groups.

**Figure 4.**
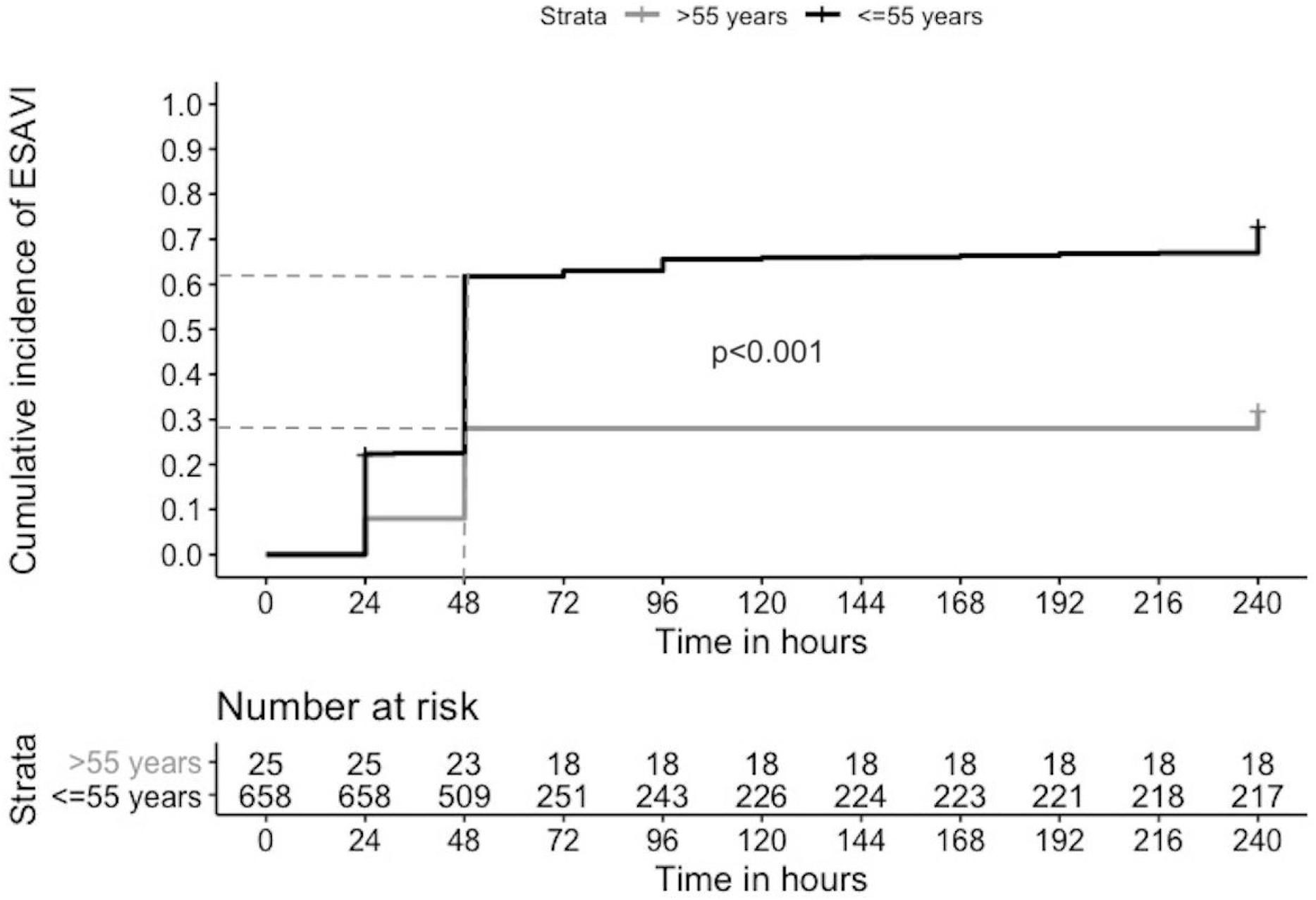
Incidence of Events Supposedly Attributed to Vaccines and Immunizations (ESAVI) according to age. Kaplan Meier curve. The dotted line corresponds to the estimated incidence of ESAVI at 48 hours. Number at risk is the number of people at risk every 24 hours from the date of vaccination. The p value compares the curves between those over 55 and up to 55 years.

Both age and sex were factors associated with the incidence of ESAVI regardless of the conditions prior to vaccination. **Table 2**

**Table 2.** Multivariate Cox regression analysis. Factors associated with the Incidence of Events Supposedly Attributed to Vaccines and Immunizations (ESAVI).

| Variables | HRcr | CI95% | p value | HRadj | CI95% | p value |
| --- | --- | --- | --- | --- | --- | --- |
| Age up to 55 años | 2.88 | 1.43-5.80 | 0.003 | 2.66 | 1.32-5.38 | 0.006 |
| Female | 1.41 | 1.15-1.72 | 0.001 | 1.38 | 1.13-1.68 | 0.002 |
| Family history of reaction to vaccines | 0.82 | 0.30-2.19 | 0.689 | 0.88 | 0.33-2.37 | 0.809 |
| Allergy prior to vaccination | 1.16 | 0.80-1.70 | 0.423 | 1.14 | 0.78-1.66 | 0.499 |
| Diabetes Mellitus | 0.71 | 0.23-2.22 | 0.561 | 0.82 | 0.26-2.56 | 0.736 |
| Any medical condition prior to vaccination | 1.22 | 0.82-1.81 | 0.320 | 1.22 | 0.82-1.82 | 0.319 |

HRcr: crude hazard ratio CI 95%: confidence interval 95% HRaj: adjusted hazard ratio

## Discussion

In our study, the incidence of events supposedly attributed to the Sputnik V vaccine among the first 707 vaccinated health workers was 71.3%; it was 17 times higher than that reported of 4.7% by the “5th report of surveillance of the safety of vaccines of the National Ministry of Health” ^7^ and similar to that reported in the interim analysis of russian COVID-19 phase 3 vaccine (Sputnik) trial (64.7%).^9^ Only one person (0.1%) required hospitalization without complications in our study, and 45 (0.3%) in phase 3 Sputnik study.^9^

The incidence for pain at the injection site, fever and headache in our study were on the range reported by Sputnik V vaccine studies.^2,9^ However, in our study we found muscular pain reported more than twice than in phase 1 y 2 Sputnik vaccine study (58% vs 23% respectively);^2^ no data on muscular pain was reported in the interim analysis of russian COVID-19 phase 3 vaccine (Sputnik) trial.^9^

In our study, in concordance with the study on the vaccine BNT162b2 mRNA Covid-19 (Pfizer)^8^ a higher incidence of ESAVI was observed in people aged up to 55 years. Adverse events among older than 55 were 71% for Pfizer vaccine compared to 32% in our study, and in younger than 55 years old it was 83% vs 72.8% respectively.^8^ In the recent publication of the interim analysis of russian vaccine, they found a higher presence of antibodies specific to the receptor-binding domain of SARS-CoV-2 glycoprotein S among younger participants compared to the older ones but they did not make comparisons of ESAVI between age groups in participants who received the vaccine to understand reactogenicity related to adaptive immunogenicity response.^9^

We also found a higher incidence of ESAVI among women compared to men (65% vs 50% respectively), we did not find comparison of ESAVI by sex in other studies.

Symptoms that appeared shortly after vaccination are usually mild and self-limited and rarely have serious consequences. However to understand the frequency and kind of symptoms attributable to vaccination is important to understand reactogenicity.^10^ The reactogenicity refers to a subset or reaction that occurred shortly after vaccination considered as physical signs of the inflammatory response to the vaccine.^11^ Reactogenicity may contribute to the predisposition to vaccination. A person could perceive a vaccine as too much reactogenic thus could reject additional doses or the health professional could take the option of not recommending it. Maintaining high vaccine coverage is critical to the success of vaccination programs.^12^

On the other hand, an association between reactogenicity and adaptive responses with early innate responses is described, but a predictive association between reactogenicity and the adaptive response was not demonstrated, lacking evidence on the known concept of “no local pain, no gain in immunity”.^13^

Recently Sadoff y cols., show preliminary result of the Phase 1 and 2 multicenter, randomized, double-blind, placebo-controlled clinical trial of the vaccine Ad26.COV2.S (Johnson & Johnson) with 805 healthy volunteers in two age cohort to assess the safety, reactogenicity and immunogenicity.^14^ In accordance to this study, our investigation showed, after the first doses, a trend to a decrease in the incidence of adverse effect with the increase of age. Local and systemic reaction reactions occurred on the day of immunization or the next day and generally resolved within 24 hours.

Systemic reactions were largely responsive to antipyretic drugs and the need for prophylactic use of these drugs was not identified. After the second dose among participants between the ages of 18 and 55, the incidence of systemic adverse events was much lower than after the first immunization, regardless of having used low or high dose, a finding that contrasts with observations made to messenger RNA-based vaccines, for which the second dose has been associated with increased reactogenicity.^15^

It is worth mentioning what was observed in the work of the University of Oxford with the AstraZeneca vaccine, a subgroup inadvertently received half a dose of the first vaccine and the second full dose. After this, this subgroup was followed up, and not only it was observed less reactogenicity but also higher immunogenicity.^15,16^

The response rate in our study was very high, in part because the ESAVI collection tool was explained at the time of vaccination by an epidemiologist, and reinforced by telephone, by a nurse. On the other hand, the origin of the vaccine and the delayed publication of phase 3 questioned by the media could influence a greater report. The tool used (self-report form) is being analyzed in its validity and reliability.

The continuity of this line of research includes a study, in the field of social epidemiology, about the social representations of prevention, in particular the “biopoliticization” of the epidemic. Also epidemiological surveillance on the incidence of adverse effects long-term and determination of the immune response in relation to reactogenicity at 72 hours are ongoing. Even though there is evidence that the immune response is lower in older adults and in men ^17,18^, understanding whether the efficacy of the vaccine is lower in people with low reactogenicity is useful to define the vaccination scheme.

## Data Availability

All raw data is available under request.

